# Indicators of improved emotion behaviour in 6–14-year-old children following a 4-week placebo controlled prebiotic supplement intervention at home with a parent

**DOI:** 10.1101/2024.10.07.24314997

**Authors:** Nicola Johnstone, Kathrin Cohen Kadosh

## Abstract

In this double-blind placebo-controlled randomised intervention we investigated the potential benefits of a prebiotic supplement on children’s well-being in a home setting. The primary aim was to determine if this supplement could effectively reduce anxiety, improve mood, and enhance cognitive function, similar to findings in young adults.

Fifty-three healthy children, aged 6 to 14, participated in an 8-week trial. The trial consisted of three testing time points; day zero marked the baseline measurement (T1) followed by a 28-day supplement intervention period during which they consumed 5.5 grams of galactooligosaccharides (GOS) daily under parental guidance. Endline measures (T2) were conducted on the last day of supplement consumption, with a final follow-up testing session (T3) on day 56. Primary outcomes were trait anxiety using a questionnaire and emotional behavior in a dot-probe task on responses to positive and negative images. Secondary outcomes encompassed depression levels, cognitive function tests, and dietary intake recorded in a 4-day food diary. Additionally, we explored whether parents’ emotional behavior had an impact on children’s responses.

While our statistical analysis did not reveal significant effects of GOS, there were noteworthy trends. Trait anxiety levels decreased over time in both groups, with a more pronounced decrease in the GOS group. The GOS group exhibited reduced negative emotional responses compared to the placebo group, and post-trial depression levels decreased in the GOS group over time. Although parental emotional responses correlated with various emotional outcomes in children, they did not influence the intervention effects. In conclusion, these findings suggest positive trends in line with our hypotheses however further investigation with greater statistical power would be beneficial.

---

Prebiotics are psychobiotic compounds that exert improvements in emotional well-being or cognition via the gut microbiome ^1^. Although these compounds have demonstrated potential in improving the health and well-being of adults, the evidence regarding their benefits for children and young people (CYP) is inconsistent ^2,3^. This presents a significant challenge, particularly as mental health issues among CYP are on the rise ^4–6^. This not only places a substantial burden on healthcare services, but also impacts the affected young individuals and their caregivers. Additionally, unresolved mental difficulties before the age of 14 can significantly increase the risk of lifelong challenges for individuals ^7^.

Securing effective mental health interventions for CYP remains a persistent challenge, characterized by delays and limited effectiveness ^8^. Additionally, the stigma surrounding mental health issues often deter CYP from seeking help. A recent research review underscores an imbalance in the literature, with an overemphasis on clinical populations and disregards the stigma associated with problematic mental health in young people ^9^. This research gap emphasized the importance of adapting successful strategies to support subclinical CYP, aiming to prevent the development of detrimental coping mechanisms or maladaptive behaviours. Therefore, it is crucial to prioritize early intervention strategies for subclinical groups rooted in health-oriented principles, rather than solely disease reduction. Our previous work with young individuals facing mental health challenges revealed their desire for autonomy in maintaining well-being, coupled with a need for evidence-backed guidance ^2^. Consequently, the validation of accessible interventions, such as psychobiotics, becomes paramount in easing the burden of poor mental health.

This study aimed to investigate the potential of a prebiotic to enhance emotional well-being among CYP aged 6-14 in a home-based setting. In our recent study involving females in late adolescence/emerging adulthood, we observed anxiolytic effects, improved emotion behaviours and changes in the gut microbiome composition after consumption of a prebiotic supplement containing GOS, which induced changes in the gut microbiome composition ^10^. Further, there are compelling arguments in favour of targeting the gut microbiome therapeutically in adolescence ^11,12^. Notably, existing studies on psychobiotic interventions in CYP have predominantly centred around clinical populations ^13,14^, cognitive challenges ^15^, or neurodevelopmental conditions ^16–19^. Considering this, we aimed to extend our successful testing protocol to a younger, typically developing cohort. The ultimate objective is to develop a readily applicable and widely accessible dietary intervention (e.g., a prebiotic) to promote adaptive emotional behaviors. This could potentially pave the way for its incorporation in settings like school meals.

To evaluate the practicality and impact of our intervention, we conducted a randomised, double-blind, fully remote, placebo-controlled trial involving GOS supplementation. This approach was designed to minimize researcher involvement, thereby mitigating potential performance effects or biases. Our investigation specifically focused on whether GOS intake enhances emotional behavior or cognitive processing, utilizing behavioral tasks and psychological testing in healthy participants aged 6-14 years. The primary outcomes under consideration were trait anxiety and emotional bias. Our hypothesis posited that GOS supplementation would result in reduced trait anxiety levels and emotional bias to negative emotions in a dot-probe task. Secondary outcomes were depression levels, cognitive function of attention, and nutritional intake. Drawing on previous findings in young adults, we anticipated a decrease in depression levels in the GOS supplement group and improved efficiency in the attentional networks, as measured by the attention network test. Additionally, we hypothesised a reduction in habitual dietary sugar intake due to GOS supplementation. Tertiary outcomes explored the influence of parent-measured emotional behaviour on child outcomes. As part of exploratory analyses, we sought to understand the impact of parental emotion behaviours on child outcomes and ascertain their strength. All outcomes were assessed for intervention effects and potential longevity during a follow-up period without dietary intervention.

## Method

This study received favourable ethical opinion from the University of Surrey ethics committee. Participating parents provided written informed consent as parents/guardians and for individual participation. Children provided assent prior to study participation. Participants were compensated for their time (£50). This protocol was registered on https://clinicaltrials.gov/ [NCT06258135] on February 6, 2024.

### Participants and design

This two-arm, randomised, double-blind placebo-controlled trial aimed to recruit 60 healthy children aged 6-14 years with a parent between March 2021 and November 2021. Based on a previous anxiolytic placebo controlled double blind trial of a GOS dietary intervention from our lab ^10^, we estimated a sample size of 30 per group was required to replicate effects in a younger group. Siblings from the same family could participate with a single parent. Eligibility criteria was based on child characteristics, namely aged 6-14 years old with no current clinical levels of anxiety and/or co-morbid psychological diagnoses, no developmental disorders, no gastro-intestinal problems or disease, and no restrictive diets (e.g., lactose intolerance, gluten free). Recruitment was via word of mouth and invitations were extended to our existing participant databases. 57 children were screened for eligibility, 1 was excluded due to dietary restrictions. 56 children were eligible to participate (from 37 separate families) and 3 declined to take part. 53 children enrolled in the study. Once enrolled, children were randomised to the GOS or placebo group 1:1 familywise (so siblings received the same supplement to maintain allocation concealment), 28 children were allocated to the GOS group, and 25 to the placebo group. Supplements were supplied and blinded at packaging by FrieslandCampina, NL. The GOS and placebo supplements were provided in powdered form in coded sachets 7.5 g/day (corresponding with 5.5 g/day active Biotis® GOS (FrieslandCampina, Amersfoort, the Netherlands) and 7.5 g/day Maltodextrin Maltosweet G 181 (Tate & Lyle) as placebo. The taste of both supplements can be characterized as slightly sweet. Parents/caregivers were instructed to assist their children in remembering to consume the supplement each day with breakfast. Participating families were advised to maintain their habitual diets for the duration of the trial.

Outcomes were assessed prior to intervention (baseline, T1), at the end of the 4-week dietary intervention (endline, T2) and 4 weeks later (follow up, T3). Primary outcomes specified that GOS would induce and sustain improved emotion ability over the assessment period evidenced by lower anxiety levels as measured by the trait subscale of the state-trait anxiety inventory ^20^, and improved emotion behaviour as measured by the dot-probe task. Secondary outcomes similarly specified improved depression levels and cognitive function as measured by the child depression inventory ^21^ and attention network test ^22^. Changes in habitual nutritional intake were also assessed. Tertiary outcomes examined the influence of parental emotion behaviour on child outcomes over the assessment period.

### Procedure

For this project, we adapted a previously successful supplement intervention protocol ^10^ for at-home use (Figure 1). A 28 day supply of supplements was mailed to the participants homes, with a link to an online testing platform hosting the study tasks ^23,24^. For baseline measurements, participants (children and parents) were instructed to complete the tasks online (questionnaires and behavioural tasks) and the following day children were to begin consuming one supplement per day for 28 consecutive days. For the first 4 days of supplement consumption, a food diary was kept by both parents and children and again for the final 4 days of the supplement consumption, and again 28 days later. Outcomes were assessed at the end of dietary intervention (T2) and 28 days later (T3).

**Figure 1.**
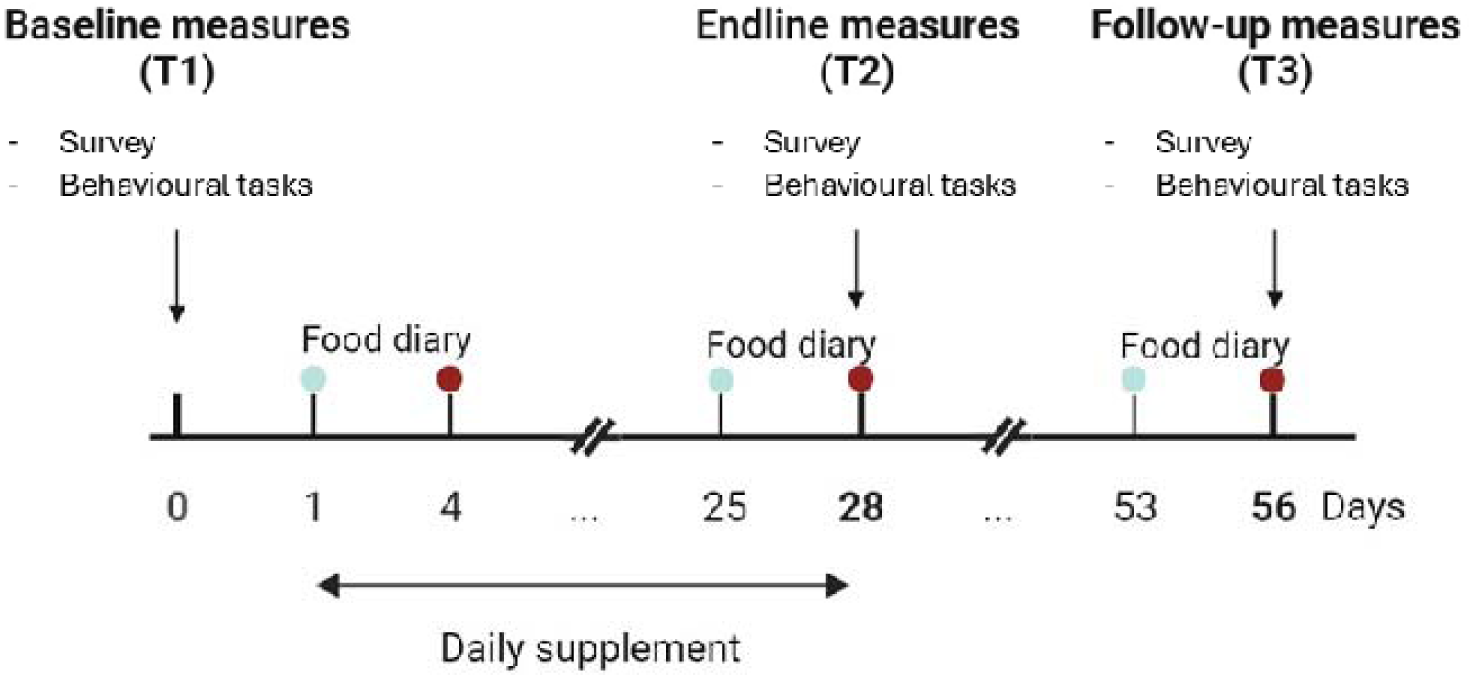
The study protocol outlines the timing of the three testing points spanning 56 days. Day Zero marks the baseline measurement (T1) followed by a 28-day supplement intervention. Endline measures (T2) were conducted on the last day of supplement consumption, with a final follow-up testing session (T3) around day 56. The duration for recording food diaries is also marked. All measures were administered through online platforms and included a survey with validated questionnaires measuring trait anxiety, depression, and additional demographic variables. Additionally, two behavioural tasks were employed to measure emotion behaviour and cognitive processing.

The flow of participants is outlined in Figure 2. Of the 53 children enrolled, 11 children did not complete T1 outcomes. 42 children returned data at T1, thereafter, 2 children requested data withdrawn. 8 children declined to complete T2 measures, thus 32 children returned data at T2 (with n = 17 for the GOS group and n= 15 for the placebo group). 3 children declined to complete T3 measures resulting in 29 children returning data at T3. Of the 40 children with T1 data available for analysis, 21 had been allocated to the intervention group (14 females, *M*_age_ = 11.2 years, *SD =* 2.91) and 19 had been allocated to the placebo group (11 females, *M*_age_ = 12.7 years, *SD =* 1.06).

**Figure 2.**
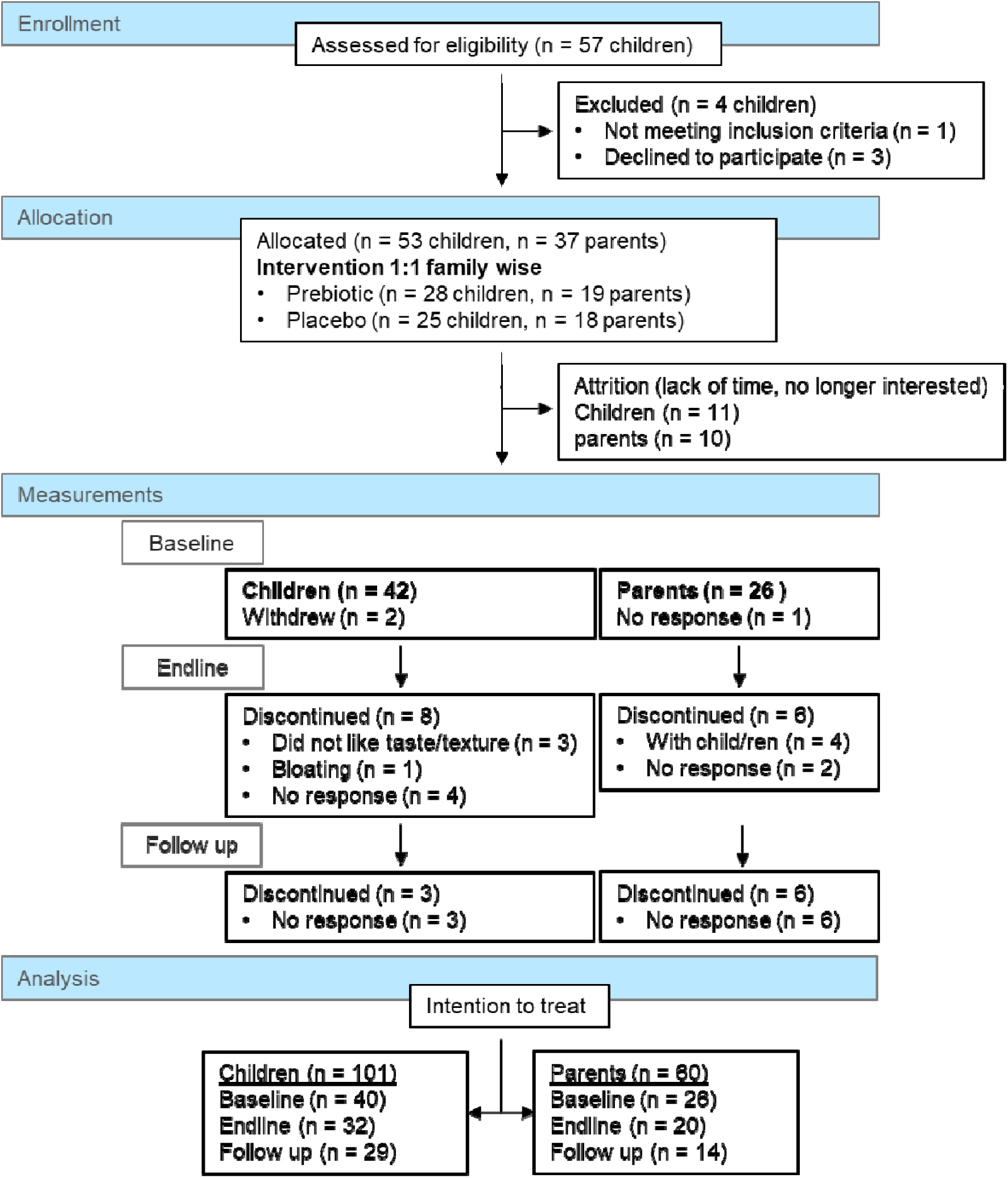
Participant flow chart showing the number of participants (children and their parents) progressing through the study milestones.

### Protocol adherence

Participants reported the date supplements were started, and approximately 3 weeks after this were sent a reminder to complete the T2 assessments on day 28. Participants confirmed that the course of supplements would be complete on that date or stated an alternative date for T2 completion (due to missing a day, beginning supplementation later than reported, or family availability). 3 weeks after T2 completion, participants were sent another reminder to complete T3 assessments on a specified date (28 days after T2). Participants who did not engage were re-contacted periodically for a duration of 2 months after last contact, then presumed to have discontinued. Average time between T1 and T2 was 31.5 days (*SD* = 3.9; range 24-39 days), and between T2 and T3 37 days (*SD* = 16.8, range 16-80 days).

### Sample Attrition

In total 4 children explicitly discontinued participation. Parents explained that this was because their children did not like the taste or texture of the supplement, or that they did not like how it affected their food (n = 3) or cited bloating (n = 1; as per the product information leaflet). The remaining attrition (11 children in total) was due to lost contact; 8 children at T2, and 3 children at T3. For those who ceased participation descriptive demographic and outcome variables (collapsed across time) were similar to those who continued participation, e.g., age; GOS group *M* = 12.9 (*SD =* 1.38), placebo group *M* = 11.88 (*SD =* 1.36); trait anxiety; GOS group *M* = 31.17 (*SD =* 7.73), placebo group *M* = 37.83 (*SD =* 9.41).

### Child measures

**Trait anxiety** was measured using the trait subscale from the state-trait anxiety measure for children ^20^ (STAIC). The STAIC consists of two 20-item scales that measure state and trait anxiety in children. The trait scale measures longer-term trait anxiety, which addresses how the child generally feels, whereas the state anxiety scale measures current anxiety state in the child. Children chose one of three responses (hardly = 1, sometimes = 2, often = 3) to a statement e.g., “*I feel unhappy…*”. Items were summed, with greater values reflecting greater anxiety. Reliability was calculated on 20 items at each time point and was found to be good (baseline α = 0.916, endline α = 0.917, follow up α = 0.923). Scores on the trait scale range from 20 – 60, with US sample norms around *M =* 35.7, *SD* = 6.6 (collapsed across males and females).

**Emotion processing** was measured using the dot-probe ^25^ task on their home laptop or computer, delivered via an experimental software ^24^ hosted online at Pavlovia (https://pavlovia.org/). The dot-probe task assesses implicit attentional biases towards emotion information, such as emotional expressions. During the dot-probe task participants were instructed to focus on a central fixation cross. After an interval, two pictures of faces with emotional expressions appeared on both sides of the screen. The faces were a combination of angry expressions (to draw out negative emotions), happy expressions (to draw out positive emotions) and calm expressions (for non-emotional response). Each emotional expression always appeared with a calm expression. The faces were visible for 500 ms, before a dot is presented in the location of one former picture. Children had to use the arrow keys to indicate if the dot was on the left or right side of the screen. The response time (RT) to the dot was recorded. *Positive vigilance* to happy faces and *negative vigilance* to angry faces was calculated from the RTs of correct responses by subtracting congruent RTs (e.g., dot in the same location as emotional face) from incongruent RTs (e.g. dot in opposite location from emotional face) for happy and angry faces separately.

**Depression** was measured using the child depression inventory ^21^ (CDI). The CDI is a 27-item measure of unidimensional depressive symptoms in children. Children are presented with groups of three sentences and asked to choose the one that best describes him or her in the past two weeks. Sentences are scored a = 0; b = 1; c = 2 with some items reversed, and totals are summed. Scales range from 0 to 54, and higher scores indicate greater levels of depressive symptoms. Reliability was calculated on 26 items at each time point and was found to be good (baseline α = 0.914, endline α = 0.934, follow up α = 0.96). In school aged children who are not depressed, mean scores on the CDI are around 8-10, with standard deviations of 7-8. Clinical levels of depression are indicated in those scoring above 17-19 points.

**The attention network test** (ANT) adapted for use by children ^22^ was used to measure cognition, at home using the online software as for the dot-probe task. The ANT assesses performance in three key attention networks: the alerting, orienting and executive function network. The ANT combines cued reaction time trials with flanker task trials to elicit varying reaction times that represent efficiency in three distinct processes of attention; alerting, orienting and executive control ^26^. There are 3 blocks of 48 trials (after a 15-trial practice block with performance feedback). Trials were comprised of a fixation cross of duration jittered between 400-1600 ms, followed by a warning cue presented for 100 ms, another fixation cross of 100 ms, then target/flanker combinations for a maximum 1700 ms, after which if no response was made the next trial began or in the instance of a response a post fixation period occurred until 4000 ms passed from trial onset. There were 3 different conditions (neutral, congruent, and incongruent) and 4 different cue conditions (spatial, double, none or center) resulting in 12 different trial types, each of which was presented 12 times across the 3 blocks. Alerting and orienting networks were estimated based on reaction rate (inverse transform on individual trials of reaction time, to estimate speed of response) following the four distinct cue trials. *Alerting network* efficiency was computed from median reaction rate in no cue condition minus median reaction rate in double cue condition. *Orienting* efficiency was calculated as median reaction rate in center cue condition minus median reaction rate in spatial cue condition. *Executive function* efficiency was computed from target types; median reaction rate to trails with target incongruent to flanker stimuli minus the median reaction rate to trials where target was congruent with flanker stimuli. Responses quicker than 200 ms (too quick) were re-coded as errors and not included in efficiency analysis. Missed responses (too slow) were re-coded as errors. The median number of errors across the 12 different trial types, participants, and time was 2, with no difference dependent on trial type. Only participants who accurately responded to at least 8/12 (e.g., clearly greater than chance) trials on each of the 12-trial condition/cue combinations were included in the efficiency analysis. On this basis, one participant was not included in the endline efficiency analysis.

**Nutritional intake** was measured using a food diary over 4 days. Parents were asked to help children record everything they ate and drank, including brands, portion sizes and time of consumption. These data were entered into a nutritional analysis database ^27^ that estimated daily energy intake, and macronutrient intakes that were then averaged over 4 days. The reported macronutrients reported were converted to a percentage of average energy intake using 4 calories per gram for carbohydrates and protein, 9 calories per gram for fat and 8 calories per gram for alcohol. Sugars are calculated from mono- and disaccharides (excluding oligosaccharides) and free sugars encompass all added sugars, including those from fruit juice and honey.

**Demographic** measures were collected by self-report, height in centimeters and weight in kilograms and sex. Age was calculated from reported date of birth and testing date. Pubertal maturation was measured using the Petersen development scale ^28^ (PDS). The PDS is a self-report 7 item instrument, each item has four statements (scored 1-4) with one statement selected per item that corresponds with current stage of development. Items are averaged to give a development score; higher scores indicate greater maturation.

### Parental measures

Parents completed the same dot-probe task, ANT task, food diary and demographic measures in addition to adult equivalent measures of trait anxiety ^29^ (STAI) and depression ^30^ (Becks Depression Inventory, BDI). The trait sub-scale of the STAI has 20 items for assessing trait anxiety. Trait anxiety items include: “I worry too much over something that really doesn’t matter” and “I am content; I am a steady person.” All items are rated on a 4-point (1-4) scale (e.g., from “Almost Never” to “Almost Always”). Score range is from 20-80 and higher scores indicate greater anxiety. Reliability was good calculated on 20 items at each time point (baseline α = 0.915, endline α = 0.932, follow up α = 0.934). The BDI is a unidimensional measure of depression symptoms of 21 items using a four-point scale which ranges from 0 (symptom not present) to 3 (symptom very intense). Values are summed, score range is from 0-84 and greater values indicate more depression symptoms. Reliability was good at each time point (21 items, baseline α = 0.789, endline α = 0.848, follow up α = 0.789).

### Statistical analysis

The statistical analysis plan was written for intention to treat analysis using all available data and performed in R ^31^. Missing data was not imputed. For all outcomes we planned to use linear mixed models to estimate fixed effects of the Supplement group (GOS or placebo) and time period (intervention effects for contrast T1 – T2; and longevity which contrasts T1-T3). Random effects were specified at the participant level. Sex, age, and pubertal maturation were investigated as covariates. For analysis of nutritional intake, the children’s BMI centile was investigated as a covariate. For tertiary child outcomes, parent emotion-related measures were investigated as covariates (trait anxiety, depression, negative vigilance, positive vigilance). Restricted Maximum Likelihood (REML) was used to estimate variance components. To show that GOS influenced an outcome in contrast to the placebo group, we examined interactions of supplement group during the intervention period. To show longevity of effects, we examined the interaction of supplement group during the follow up period. Outcomes are significant at *p* < .05 and considered trend level *p* < .1.

### Post-hoc analysis

We altered the planned statistical analysis following the study completion due to disrupted data collection. Unforeseen circumstances affected the platform hosting our online tasks that resulted in participants being unable to complete the tasks at the correct time. While this was quickly resolved, there remained ongoing related complications that did not become apparent until data collection ceased. This chiefly affected follow up data (T3) thus for emotion behaviour and cognitive processing we assessed only the intervention period (T1-T2) using ANCOVA models. Similarly, we assessed only the intervention period with ANCOVA for nutritional outcomes. This was due to a low return rate of the food diaries across the study, and particularly at follow up with participant attrition. We speculate that the remote nature of the task lessened the focus on completing the food diaries, given that both survey and computer tasks were presented together, and were not so laborious for parents who supported children with food diary completion.

## Results

### Descriptive statistics

Of the 53 children enrolled, 40 provided baseline measures (21 in GOS group and 19 in placebo group). Participating children’s demographic details are described in Table 1.

**Table 1.** Demographic measures at baseline for GOS and placebo groups split by male and female children.

|  | GOS mean (SD) |  |  | Placebo mean (SD) |  |  |
| --- | --- | --- | --- | --- | --- | --- |
|  | Female | Male | Total | Female | Male | Total |
| Sex (n) | 14 | 7 | 21 | 11 | 8 | 19 |
| Age (years, range 6-14) | 11.2 (2.91) | 11.6 (2.19) | 11.61 (2.52) | 12.7 (1.06) | 11.8 (1.90) | 12.32 (1.49) |
| Pubertal maturation (score range 1-4) | 2.26 (.081) | 1.79 (0.67) | 2.10 (0.78) | 2.29 (0.46) | 1.87 (0.47) | 2.12 (0.50) |
| Weight (kg) | 44.6 (17.2) | 36.5 (13.6) | 41.89 (16.17) | 45.3 (18.8) | 40.4 (4.97) | 43.11 (14.59) |
| Height (cm) | 143 (18.2) | 142 (23.2) | 142.48 (19.39) | 156 (9.83) | 157 (16.4) | 156.56 (12.72) |
| BMI Centile (%) | 72.5 (23.7) | 59.8 (30.7) | 69.1 (25.2) | 41.6 (37.5) | 46.3 (32.1) | 44.2 (32.9) |

### Primary outcomes

We anticipated that GOS consumption, as opposed to placebo consumption, would lead to a decrease in trait anxiety. Baseline trait anxiety scores were similar with no group difference (*M*_GOS_ = 34.27, *SD* = 7.58, *M*_placebo_ = 37.05, *SD* = 10.24, *p* = 0.335). Trait anxiety scores were normally distributed, and there were no significant covariates from sex, age, or pubertal maturation. The mixed model identified a reduction in trait anxiety scores following intervention period (*p* = .048) and in the follow up period (*p* = .016) but no interaction of time and supplement group. Simple effects analysis found the reduction in trait anxiety scores was greater in the GOS group (intervention period, T2-T1; β −2.68 points, *SE* = 1.33, *p* = .090, follow up period, T3-T1; β −3.37 points, *SE* = 1.36, *p* = .031) than in the placebo group (intervention period, T2-T1; β −2.69 points, *SE* = 1.46, *p* = .131, follow up period, T3-T1; β −2.59 points, *SE* = 1.51, *p* = .166), Figure 3A.

**Figure 3:**
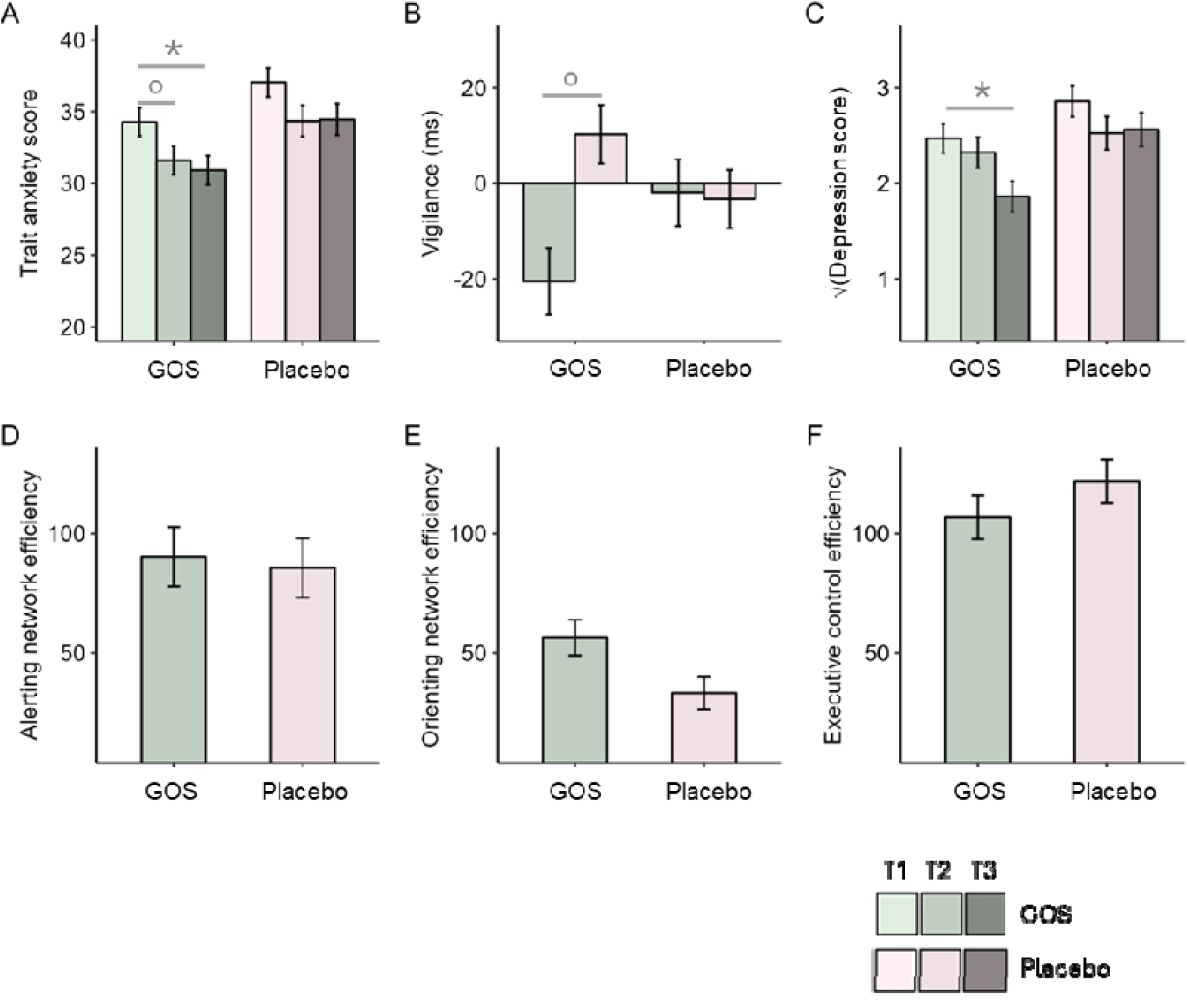
Statistical analysis plots for emotion-related and cognitive outcomes. Bars in green represent the GOS group outcomes, while bars in pink depict the placebo group outcomes; shades of both colours darken with each testing point. **Panel A** presents estimated marginal means (EMM) for trait anxiety assessed by linear mixed modelling. Simple effects analysis compares T2 and T3 responses to T1 within GOS and placebo groups separately. In the GOS group, T2 anxiety decreased, and by T3, a significant reduction in trait anxiety was observed. **Panel B** displays EMM for the secondary outcome of emotion behavior during the dot probe task at T2. ANCOVA analysis reveals decreased vigilance to negative information in the GOS group compared to the placebo group. **Panel C** illustrates square root transformed EMM for the secondary outcome of depression, as assessed by linear mixed modelling. Simple effects analysis compares T2 and T3 responses to T1 within GOS and placebo groups separately. In the GOS group, T3 depression was reduced compared to T1, with no such change observed in the placebo group. **Panels D, E, and F** present cognitive outcomes with EEM plotted following ANCOVA. While no significant effects were observed, it is noteworthy that the GOS group demonstrated more efficient results in all three networks compared to the placebo group. Error bars are standard errors of EMM. Significance levels: * p < .05, LJ p < .10.

On **emotion processing** an increase in positive vigilance to happy faces and/ or reduction in negative vigilance to angry faces is desired. For this analysis, we proceeded with an Ancova of the endline measures, with baselines as a covariate due to obtaining only for 5 children (GOS group) and for 8 children (placebo group) completed datasets at follow up. Baseline vigilance scores did not differ between groups for negative (*p* = .323) or positive (*p* = 0.555) emotions, and there was no influence of sex, age, or pubertal maturational score. Variables were normally distributed. There was no group effect following intervention in an ancova model considering positive and negative vigilance simultaneously, however, simple effects contrasts of group responses to negative and positive vigilance separately showed a trend towards a reduction in negative vigilance following intervention in the GOS group compared to placebo group (β −30.75 ms, *SE* = 18.58, *p* = .105), Figure 3B.

### Secondary outcomes

On **depression** a reduction in scores was predicted in the GOS group compared to the placebo group. Distribution of scores on the CDI instrument are right skewed with a high incidence of zeros. This was reflected in the baseline scores (*M*_GOS_ = 8.11, *SD* = 4.62, *M*_placebo_ = 12.00, *SD* = 10.02). The square root transform normalised the distribution and resultantly there was no baseline group difference, *p* = 0.391. There were no significant covariates from sex, age, or pubertal maturation. The mixed model identified a reduction in depression levels in the follow up period (*p* = .008) but no interaction of time and group. Simple effects analysis found the reduction in depression levels was greater in the GOS group during follow up (T3-T1; β −0.61, *SE* = 0.22, *p* = .015), Figure 3C.

On **cognitive processing**, we evaluated networks involved in alerting attention, orienting attention, and executive function. In alerting and orienting, desired improvements in efficiency are illustrated by increased processing times (higher numbers). In executive function, improved efficiency is represented by reductions in processing times (smaller numbers). In all three networks, there was no group differences at baseline (alerting, *p =* 0.360; orienting *p* = 0.544; executive function, *p* = 0.672) and all efficiency outcomes were normally distributed. There was no covariate influence on any network from sex, age, or pubertal maturation. To test supplement intervention effects, ancova models were used with endline measures as outcomes and baseline measures as covariates as there were only datasets of 6 children (GOS group) and of 5 children (placebo group) available at follow up. For alerting efficiency, there was no effect of supplement group (*p* = 0.895), there was no supplement group effect for the orienting network (*p* = 0.268), or for executive function (*p* = 0.581). Numerically, at T2 all networks appeared more efficient for the GOS group (e.g., responding in the expected direction) compared to the placebo group (Figure 3D-F).

On **nutrient intake**, we have previously found reduced intakes of carbohydrate, sugars and increased fat intake following GOS supplementation ^32^ . We anticipated similar results here. In the GOS group, 13 food diaries were returned at baseline, 14 at endline, and 6 at follow up. In the placebo group, there were 10 at baseline, 9 at endline and 8 at follow up. To conserve power, we assessed only endline group effects with baseline measures as covariates. A descriptive overview of baseline energy intake and key nutrients is presented in Table 2. Table 3 presents the outcome of the ancova analyses. Free sugar differed following intervention, where the placebo group had lower reported intakes than the GOS group, where there had been at similar levels at baseline. Monounsaturated fat also differed following intervention, where the placebo group had lower reported intakes than the GOS group, where there had been at similar levels at baseline.

**Table 2.** Descriptive overview of nutritional elements analysed at baseline. Data are the average of 4-day comprehensive food diaries.

| Intake | GOS |  | Placebo |  |
| --- | --- | --- | --- | --- |
|  | Child, N = 13<sup>1</sup> | Parent, N = 10<sup>1</sup> | Child, N = 10<sup>1</sup> | Parent, N = 8<sup>1</sup> |
| Energy (kcal) | 1,737.80<br>(305.82) | 1,604.34 (325.89) | 1,894.48<br>(487.39) | 1,775.83<br>(460.07) |
| Carbohydrate (%) | 49.75 (4.41) | 44.04 (6.80) | 47.70 (4.34) | 43.27 (6.43) |
| Fat (%) | 33.58 (4.39) | 34.64 (4.20) | 36.16 (4.17) | 36.08 (3.28) |
| Protein (%) | 14.80 (2.28) | 16.36 (4.31) | 14.55 (3.31) | 16.51 (4.02) |
| Fibre (g) | 20.81 (5.68) | 17.97 (5.74) | 21.98 (8.18) | 23.29 (6.47) |
| Sugars (%) | 19.06 (3.94) | 18.04 (2.24) | 16.36 (3.82) | 13.33 (5.02) |
| Free sugar (%) | 6.45 (3.49) | 7.37 (2.47) | 7.13 (4.84) | 4.46 (2.68) |
| Saturated fat (%) | 12.99 (2.43) | 13.01 (2.53) | 13.75 (3.89) | 11.28 (2.40) |
| Monounsaturated fat (%) | 9.69 (1.43) | 12.02 (2.02) | 10.10 (1.97) | 10.68 (2.87) |
| Diet |  |  |  |  |
| Omnivore | 11 / 13<br>(85%) |  | 6 / 10 (60%) |  |
| Vegetarian | 2 / 13<br>(15%) |  | 4 / 10 (40%) |  |

<sup>1</sup>Mean (SD); % = percentage of energy intake.

**Table 3.** Statistical outcomes of intervention group effects at endline assessed with ANCOVA. Estimated marginal means and standard errors are reported with significance level.

| Intake | Influential covariate | GOS |  | Placebo |  | *p* |
| --- | --- | --- | --- | --- | --- | --- |
|  |  | EMM | SE | EMM | SE |  |
| Energy (kcal.) | NA | 1456.70 | 139.44 | 1696.05 | 154.39 | 0.269 |
| Carbohydrate (%) | NA | 49.30 | 1.41 | 49.11 | 1.56 | 0.932 |
| Fat (%) | NA | 35.07 | 1.41 | 34.87 | 1.57 | 0.928 |
| Protein (%)<sup>1</sup> | Energy, Diet. | 14.11 | 0.81 | 16.07 | 0.81 | 0.101 |
| Fibre (g)<sup>2</sup> | Energy, Diet. | 19.31 | 1.10 | 19.56 | 1.10 | 0.182 |
| **Free Sugars (%)** | **NA** | **8.20** | **1.29** | **3.30** | **1.26** | **0.013** |
| Sugars (%) | NA | 18.15 | 1.06 | 16.88 | 1.18 | 0.452 |
| Saturated Fat (%) | NA | 13.87 | 0.80 | 12.91 | 0.88 | 0.429 |
| **Monounsaturated fat (%)** | **NA** | **9.38** | **0.58** | **7.46** | **0.65** | **0.041** |

<sup>1</sup> Protein as a percentage of energy intake at baseline was influenced by energy intake where greater energy consumption was associated with lower protein intake and vegetarian diets were associated with lower protein intake.
<sup>2</sup> Fibre in grams at baseline was influenced by energy intake where greater energy consumption was associated with increased fibre intake, and vegetarian diets with greater fibre intake.

### Tertiary outcomes

Parent emotion-related variables, trait anxiety, depression, negative vigilance, and positive vigilance were considered at baseline as predictors for child level outcomes. Significantly influential parent variables were included in the child outcome model to determine if parent’s emotion behaviour had an impact on the intervention or follow up period.

In terms of child trait anxiety, parents’ negative vigilance was linked to increased child anxiety (p = 0.017), while parents’ positive vigilance was associated with decreased child anxiety (p = 0.027). However, these parental vigilance factors did not affect the outcomes of the supplement intervention or the follow-up effects. Regarding child negative vigilance, parents’ negative vigilance exhibited a negative correlation (p = 0.028), whereas parents’ positive vigilance showed a positive correlation (p = 0.039). For child positive vigilance, there was no impact from parents’ emotional behavior, and no discernible effects on children’s emotional vigilance were observed following the supplement intervention. Parents’ emotional behavior did not exert any influence on child depression. In terms of child cognition within the executive control network, parents’ depression levels were positively linked to poorer child efficiency in executive control (p = 0.003). However, this association did not translate into observable effects on child responses following the supplement intervention.

In assessing nutrition outcomes, we considered the influence of the matched parental intake at baseline. If significant, this influence was included in the Ancova child outcomes model to examine the impact of parental nutritional intake on child nutritional intake. Concerning energy intake, carbohydrate, fat, protein, fibre and saturated fat, parental intake showed a positive correlation with child intake. In other words, the more of each nutrient consumed by the parent, the more consumed by the child. However, incorporating parental intakes into the child Ancova model did not affect any group effects. Notably, the intakes of free sugar, sugars and monounsaturated fat by parents were not correlated with child intakes.

## Discussion

The objective of this trial was to evaluate the potential of an easily accessible supplement intervention in enhancing the wellbeing of children and young people (CYP) by targeting emotion behaviour, cognition, or nutrient intakes. Our hypothesis was that daily supplementation with the prebiotic GOS would have anxiolytic effects, improve mood, cognition, and induce improvements in nutrient intakes. The observed results displayed positive trends aligning with our hypotheses, including a decrease in anxiety over time, reduced vigilance to negative stimuli, and decreasing depression levels in the GOS group. However, we note that these trends did not reach statistical significance when compared to the placebo group.

Previous research in children investigating the impact of prebiotics on mental health outcomes has yielded mixed or inconclusive results. This variability can be attributed to various factors such as differences in study design, population characteristics, and methodological limitations ^2,3^. Although the observed positive trends in the GOS group regarding the reduction of anxiety, negative emotional responses and depression levels in the GOS group did not reach statistical significance, they hold valuable insights and are consistent with the known effects of prebiotics observed in adult studies. However, it is essential to acknowledge the need for further validation through larger-scale studies. This is particularly crucial within in the context of typically developing children and young people, including subclinically anxious participants, who are underrepresented in the existing literature.

Growing evidence supports the use of prebiotics to enhance overall wellbeing, not only for supplements but also by incorporating prebiotic food into entire diets. In a novel 4-arm placebo controlled dietary intervention trial, which compared either followed a high prebiotic diet, or a probiotic diet or symbiotic combinations, participants on the high prebiotic diet reported improved mood over an 8-week period, especially those with initially very low fiber consumption ^33^. Another intervention focused on adults with poor dietary habits, involving a 4-week psychobiotic dietary program. The participants received educational training from a dietitian, contrasting with a generic healthy eating control group ^34^. At the conclusion of the trial, the psychobiotic diet group reported lower perceived stress scores, accompanied by changes in nutritional intake, such as increased fibre, and alternations in gut microbiome composition ^34^. Although these studies targeted adults, the focus on non-clinical populations is commendable and underscores the potential of dietary prebiotics and psychobiotics in safeguarding the well-being in CYP with persistently poor dietary habits.

For individuals already maintaining nutritionally adequate diets, the addition of prebiotic supplementation can potentially impact both nutritional intake and overall well-being. A 2-arm placebo controlled study involving GOS supplements over a 4-week period in young female adults revealed notable changes in gut microbiome composition associated with alterations in nutritional intake by the study’s conclusion ^32^. Specifically, carbohydrate and sugar intake decreased, and fat increased in the GOS group relative to the placebo group. Additionally, the GOS supplement led to higher abundances of *Bifidobacterium* which correlated with reduced carbohydrate. However, this was not replicated in the present study. We found that the placebo group exhibited lower free sugar intake at the trial’s end in comparison to the GOS group. It is essential to approach these results with caution due to the limited number of returned food diaries. The protocol’s requirement for participants, particularly parents, to maintain their food diaries might not have been efficient, potentially placing an added burden on parents in documenting both their own and their children’s food consumption. Recognizing the challenges in obtaining accurate dietary assessments from CYP (e.g., ^35^), future studies could address some of the reporting difficulties by utilizing simpler food frequency questionnaires with accurate portion sizes specifically designed for CYP, as suggested in previous research ^36^.

One noteworthy observation from the nutrition analysis is the independence of children’s intake of sugars, free sugar, and monounsaturated fats from parental intakes. Sugars are calculated from mono-and disaccharides (excluding oligosaccharides) and free sugars encompass all added sugars, including those from fruit juice and honey. As depicted in Table 2 children reported consuming more sugars and free sugars but less monounsaturated fat compared to parental reports. This variance could suggest greater autonomy among children in choosing these specific types of foods. Alternatively, it may indicate differential taste preferences between adults and children, with children showing a tendency to favour sweeter tastes over fat ^37^.

It is noteworthy that the reported intakes of free sugar fall below the World Health Organization’s recommendation of 10% of total energy intake ^38^ . However, this observation underscores the potential effectiveness of dietary interventions targeted at children, particularly those oriented towards sweet tastes and reduced fat preferences. Such interventions could play a crucial role in promoting healthier dietary habits among children.

Irrespective of the intervention effects, parent’s emotion measures exhibited a positive influence over children’s emotion measures. This impact is supported by the concept of heritability, representing the proportion of trait influenced by genetic factors. For anxiety ^39^ and depression ^40^, heritability can be substantial, increasing the likelihood that biological children will demonstrate similar tendencies. The use of self-report instruments enabled the subjective assessments of an individual’s traits within the context of their experiences and environment. This approach captures environmental sensitivity, indicating context-dependent individual reactions ^41,42^. Environmental sensitivity is bidirectionally influenced by both biological and psychosocial factors. Consequently, therapeutics interventions, such as the potential effects of prebiotics on the gut-microbiome-brain axis, offer hold promise for children (CYP) and environmentally sensitive parents by potentially balancing CNS reactivity.

In conclusion, the prebiotic intervention in CYP reveals encouraging trends in enhancing well-being using a well-tolerated supplement. However, a notable limitation of this study that should not be disregarded is the small sample size. It is crucial for future studies to address this by adequately powering trial arms to detect controlled intervention effects. This approach will enhance the robustness and reliability of findings, contributing to a more comprehensive understanding of the potential benefits of prebiotic interventions in this population.

## Data Availability

All data produced are available online at https://osf.io/nzkqw/?view_only=0f6734447fc34410908c3a20c64ba205

## Acknowledgements

This study was supported by FrieslandCampina, NL.

We thank Caroline Carvalho, Eloise Crowson, Anna Kauer, and Paul Knytl for supporting the production of research materials in this study.

## Author notes data sharing

The data that support the findings of this study are openly available in osf at https://osf.io/nzkqw/?view_only=0f6734447fc34410908c3a20c64ba205

## Authorship

KCK and NJ conceived and designed the study, acquired, analysed, and interpreted the data. Both authors wrote the manuscript and have approved the final version.

## References

1. Sarkar, A. et al. Psychobiotics and the Manipulation of Bacteria-Gut-Brain Signals. Trends Neurosci. 39, 763–781 (2016).

2. Cohen Kadosh, K., et al. Psychobiotic interventions for anxiety in young people: a systematic review and meta-analysis, with youth consultation. Transl. Psychiatry 11, 352 (2021).

3. Basso, M. et al. A systematic review of psychobiotic interventions in children and adolescents to enhance cognition functioning and emotional behaviour. Nutrients 13, 4384 (2021).

4. Pitchforth, J. et al. Mental health and well-being trends among children and young people in the UK, 1995–2014: analysis of repeated cross-sectional national health surveys. Psychol. Med. 49, 1275–1285 (2019).

5. Wright, B., Garside, M., Allgar, V., Hodkinson, R. & Thorpe, H. A large population-based study of the mental health and wellbeing of children and young people in the North of England. Clin. Child Psychol. Psychiatry 25, 877–890 (2020).

6. Cybulski, L. et al. Temporal trends in annual incidence rates for psychiatric disorders and self-harm among children and adolescents in the UK, 2003–2018. BMC Psychiatry 21, 229 (2021).

7. Mulraney, M. et al. A systematic review of the persistence of childhood mental health problems into adulthood. Neurosci. Biobehav. Rev. 129, 182–205 (2021).

8. Cotgrove, A. Editorial: The future of crisis mental health services for children and young people. Child Adolesc. Ment. Health 23, 1–3 (2018).

9. Fledderjohann, J., Erlam, J., Knowles, B. & Broadhurst, K. Mental health and care needs of British children and young people aged 6–17. Child. Youth Serv. Rev. 126, 106033 (2021).

10. Johnstone, N. et al. Anxiolytic effects of a galacto-oligosaccharides prebiotic in healthy females (18–25 years) with corresponding changes in gut bacterial composition. Sci. Rep. 11, 8302 (2021).

11. McVey Neufeld, K.-A., Luczynski, P., Seira Oriach, C., Dinan, T. G. & Cryan, J. F. What’s bugging your teen?—The microbiota and adolescent mental health. Neurosci. Biobehav. Rev. 70, 300–312 (2016).

12. McVey Neufeld, S. F., Ahn, M., Kunze, W. A. & McVey Neufeld, K.-A. Adolescence, the Microbiota-Gut-Brain Axis, and the Emergence of Psychiatric Disorders. Biol. Psychiatry (2023) doi:10.1016/j.biopsych.2023.10.006.

13. Harrison, T. M., Brown, R., Bonny, A. E., Manos, B. E. & Bravender, T. Omega-3 fatty acids and autonomic function in adolescents with anorexia: A randomized trial. Pediatr. Res. 92, 1042– 1050 (2022).

14. Manos, B. E. et al. A pilot randomized controlled trial of omega-3 fatty acid supplementation for the treatment of anxiety in adolescents with anorexia nervosa. Int. J. Eat. Disord. 51, 1367–1372 (2018).

15. Capitão, L. P. et al. Prebiotic supplementation does not affect reading and cognitive performance in children: A randomised placebo-controlled study. J. Psychopharmacol. (Oxf.) 34, 148–152 (2020).

16. Liu, Y.-W. et al. Effects of Lactobacillus plantarum PS128 on Children with Autism Spectrum Disorder in Taiwan: A Randomized, Double-Blind, Placebo-Controlled Trial. Nutrients 11, 820 (2019).

17. Billeci, L. et al. A randomized controlled trial into the effects of probiotics on electroencephalography in preschoolers with autism. Autism 27, 117–132 (2023).

18. Skott, E. et al. Effects of a synbiotic on symptoms, and daily functioning in attention deficit hyperactivity disorder – A double-blind randomized controlled trial. Brain. Behav. Immun. 89, 9– 19 (2020).

19. Ghanaatgar, M. et al. Probiotic supplement as an adjunctive therapy with Ritalin for treatment of attention-deficit hyperactivity disorder symptoms in children: a double-blind placebo-controlled randomized clinical trial. Nutr. Food Sci. 53, 19–34 (2022).

20. Spielberger, C. D. Manual for the State Trait Anxiety Inventory for Children. (Consulting Psychologists Press, Palo Alto, 1973).

21. Kovacs, M. Children’s Depression Inventory. (Multi Health Systemes Inc., New York, 1992).

22. Rueda, M. R. et al. Development of attentional networks in childhood. Neuropsychologia 42, 1029–1040 (2004).

23. Qualtrics. Qualtircs (2005).

24. Peirce, J., Hirst, R. & MacAskill, M. Building Experiments in PsychoPy. (SAGE, London, 2022).

25. Staugaard, S. R. Reliability of two versions of the dot-probe task using photographic faces. Psychol. Sci. 51, 339–350 (2009).

26. Fan, J., McCandliss, B. D., Sommer, T., Raz, A. & Posner, M. I. Testing the Efficiency and Independence of Attentional Networks. J. Cogn. Neurosci. 14, 340–347 (2002).

27. Nutritics. (2019).

28. Petersen, A. C., Crockett, L., Richards, M. & Boxer, A. A self-report measure of pubertal status: Reliability, validity, and initial norms. J. Youth Adolesc. 17, 117–133 (1988).

29. Spielberger, C. D., Gorsuch, R. L., Lushene, R., Vagg, P. R. & Jacobs, G. A. State-Trait Anxiety Inventory for Adults. (Mind Garden Inc., Palo Alto, California, 1983).

30. Beck, A. T., Steer, R. A. & Brown, G. K. BDI-II: Beck Depression Inventory Manual. (Psychological Corporation, San Antonio, Texas, 1996).

31. R Core Team. R: A Language and environment for statistical computing. R Foundation for Statistical Computing. (2023).

32. Johnstone, N. et al. Nutrient Intake and Gut Microbial Genera Changes after a 4-Week Placebo Controlled Galacto-Oligosaccharides Intervention in Young Females. Nutrients 13, 4384 (2021).

33. Freijy, T. M. et al. Effects of a high-prebiotic diet versus probiotic supplements versus synbiotics on adult mental health: The “Gut Feelings” randomised controlled trial. Front. Neurosci. 16, (2023).

34. Berding, K. et al. Feed your microbes to deal with stress: a psychobiotic diet impacts microbial stability and perceived stress in a healthy adult population. Mol. Psychiatry 28, 601–610 (2023).

35. Livingstone, M. B. E., Robson, P. J. & Wallace, J. M. W. Issues in dietary intake assessment of children and adolescents. Br. J. Nutr. 92, S213–S222 (2004).

36. Schnermann, M. E., Nöthlings, U. & Alexy, U. Empirically derived portion sizes from the DONALD study for 4 to 18 year old children and adolescents to simplify analysis of dietary data using FFQ. Public Health Nutr. 1–20 (2024) doi:10.1017/S136898002400017X.

37. Mennella, J. A., Finkbeiner, S. & Reed, D. R. The proof is in the pudding: children prefer lower fat but higher sugar than do mothers. Int. J. Obes. 36, 1285–1291 (2012).

38. Organization, W. H. Guideline: Sugars Intake for Adults and Children. (World Health Organization, 2015).

39. Purves, K. L. et al. A major role for common genetic variation in anxiety disorders. Mol. Psychiatry 25, 3292–3303 (2020).

40. Sullivan, P. F., Neale, M. C. & Kendler, K. S. Genetic Epidemiology of Major Depression: Review and Meta-Analysis. Am. J. Psychiatry 157, 1552–1562 (2000).

41. Pluess, M. Individual Differences in Environmental Sensitivity. Child Dev. Perspect. 9, 138–143 (2015).

42. Pluess, M., Lionetti, F., Aron, E. N. & Aron, A. People differ in their sensitivity to the environment: An integrated theory, measurement and empirical evidence. J. Res. Personal. 104, 104377 (2023).

